# Lifestyle Risk Factors for Cardiovascular Disease in Relation to COVID-19 Hospitalization: A Community-Based Cohort Study of 387,109 Adults in UK

**DOI:** 10.1101/2020.05.09.20096438

**Authors:** Mark Hamer, Mika Kivimäki, Catharine R. Gale, G. David Batty

## Abstract

**Aims:** It is important to identify characteristics of people who may be most at risk of COVID-19 to inform policy and intervention. Little is known about the impact of unhealthy lifestyles including smoking, physical inactivity, obesity, and excessive alcohol intake. We conducted the first large-scale general population study on lifestyle risk factors for COVID-19.

**Methods:** Prospective cohort study with national registry linkage to hospitalisation for COVID-19. Participants were 387,109 men and women (56.4 ±8.8 yr; 55.1% women) residing in England from UK Biobank study. Physical activity, smoking, and alcohol intake, were assessed by questionnaire at baseline (2006-2010). Body mass index, from measured height and weight, was used as an indicator of overall obesity. Outcome was cases of COVID-19 serious enough to warrant a hospital admission from 16-March-2020 to 26-April-2020.

**Results:** There were 760 COVID-19 cases. After adjustment for age, sex and mutually for each lifestyle factor, physical inactivity (Relative risk, 1.32, 95% confidence interval, 1.10, 1.58), smoking (1.42; 1.12, 1.79) and obesity (2.05; 1.68, 2.49) but not heavy alcohol consumption (1.12; 0.93, 1.35) were all related to COVID-19. We also found a dose-dependent increase in risk of COVID-19 with less favourable lifestyle scores, such that participants in the most adverse category had 4-fold higher risk (4.41; 2.52 - 7.71) compared to people with the most optimal lifestyle. This gradient was little affected after adjustment for a wide range of covariates. Based on UK risk factor prevalence estimates, unhealthy behaviours in combination accounted for up to 51% of the population attributable fraction of severe COVID-19.

**Conclusions and Relevance:** Our findings suggest that an unhealthy lifestyle synonymous with an elevated risk of non-communicable disease is also a risk factor for COVID-19 hospital admission, accounting for up to half of severe cases. Adopting simple lifestyle changes could lower the risk of severe infection.

## Introduction

For non-communicable disease outcomes, lifestyle risk factors have been consistently associated with morbidity, mortality and loss of disease-free years of life.^1-4^ There are also population cohort data on possible adverse effects of poor lifestyle on serious respiratory infections. For example, physical inactivity and smoking appear to be independently associated with higher risk of community-acquired pneumonia and pneumonia mortality.^5-9^ Evidence for alcohol intake and diet on risk of respiratory infection are less clear.^8,9^

A better understanding of the links between lifestyle risk factors and COVID-19 has obvious implications for prevention of severe outcomes and also in identifying characteristics of those people most at risk. We are, however, unaware of any existing data on the relation of lifestyle risk factors with COVID-19. Accordingly, we examined the association of lifestyle risk factors with new cases of COVID-19-hospitalisations in a general population-based cohort study.

## Methods

### Study Population

We used data from UK Biobank, a prospective cohort study, the sampling and procedures of which have been well described.^10^ Baseline data collection took place between 2006 and 2010 across twenty-two research assessment centres in the UK giving rise to a sample of 502,655 people aged 40 to 69 years (response rate 5.5%).^10^ Ethical approval was received from the North-West Multi-centre Research Ethics Committee, and the research was carried out in accordance with the Declaration of Helsinki of the World Medical Association, and participants gave informed consent. No specific ethical approval was required for the present analyses of anonymised data.

### Lifestyle Measures

Physical activity, smoking, and alcohol consumption were assessed by questionnaire at baseline. These characteristics have demonstrated face validity in the UK Biobank sample through their associations with mortality and cardiovascular disease.^11^ Participants were categorised into never, previous, and current smokers. From information on the weekly intake of beer and cider (1 pint =2 units), wines (1 standard glass = 2 units) and spirits (1 shot = 1 unit), we aggregated units of alcohol intake per week. Heavy alcohol intake was defined as ≥14 units in women and ≥21 units in men.^3^ Physical activity was assessed using the International Physical Activity Questionnaire (IPAQ) short form^12^ that measures duration and frequency of moderate-to-vigorous physical activity (MVPA) from all domains in the last week. Meeting activity guidelines was defined as ≥150 min/week MVPA or ≥75 min/week vigorous PA.^3^ Body weight was measured using Tanita BC418MA scales and standing height using a Seca height measure. Body mass index (BMI) was calculated [weight (kilograms)/height^2^ (meters^2^) squared] and categorised into standard groups: healthy weight < 25; overweight 25 - < 30; obese ≥ 30 kg/m^2^.

### Covariates

During the clinic visit, data were collected via self-report for ethnicity (White, South Asian, Black, Chinese, other), educational attainment (college/degree; A-level; O-level; CSEs or equivalent; National vocational Qualifications/ Higher National Diploma or equivalent; other professional qualification; none), and self-reported physician diagnosed cardiovascular diseases (heart attack, angina, stroke) and diabetes. Hypertension was defined as elevated measured blood pressure (≥140/90 mmHg) and /or use of anti-hypertensive medication.

### Ascertainment of Hospitalisation for COVID-19

Provided by Public Health England, data on COVID-19 status covered the period from 16^th^ March 2020, after which testing was restricted to those with symptoms in hospital (http://biobank.ndph.ox.ac.uk/showcase/field.cgi?id=40100). For the present analyses COVID-19 testing results up to 26^th^April 2020 were included. These data can be regarded as a proxy for hospitalisations for severe cases of the disease for England only; study members from Scotland and Wales were therefore omitted from our analytical sample. COVID-19 disease tests were performed on samples from combined nose/throat swabs, using real time polymerase chain reaction (RT-PCR) in accredited laboratories.^13^

### Statistical Analyses

Analyses were performed using SPSS Version 26. We assigned points to different levels of each lifestyle behaviour: smoking history (0=never; 1=past; 2=current), physical activity (0=meeting guidelines; 1= active but below guideline; 2=inactive), alcohol (0= moderate intake within guidelines; 1= never or very occasional; 2= heavy intake exceeding guidelines), obesity (0=healthy weight; 1=overweight; 2 = obese). Thus, scores ranged from 0 (optimal) to 8 (worst). We fitted regression models to estimate relative risk (RR) and 95% confidence intervals for associations between lifestyle scores and COVID-19. Relative risks were first adjusted for age and sex, followed by education, ethnicity, diabetes, hypertension, and cardiovascular diseases. We calculated Population Attributable Fraction (PAF) using our mutually-adjusted effect estimates on lifestyle factors and COVID-19 and lifestyle factor prevalence from Health Survey for England^14,15^ (a representative, population-based survey of adults aged 16 and over living in private households in England) to evaluate the proportion of severe COVID-19 cases that could be avoided if high-risk people adopted a healthier lifestyle:

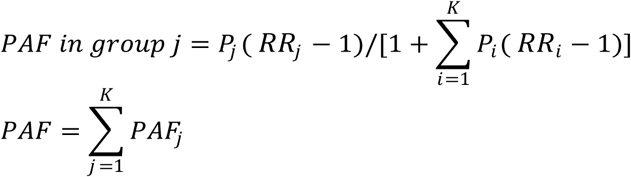

where P_i_ = proportion of the population in group i; RR_i_ = rate ratio in group i; K = number of non-reference risk groups

## Results

The analytical sample comprised 387,109 participants (56.2 ±8.0 years; 55.1% women) who were alive up to 5^th^ March 2020, and had available data on lifestyle exposures and covariates. Participants were largely white British (94.5%). Of the lifestyle factors, 33.5% exceeded alcohol intake guidelines, 23.5% were obese, 9.7% smokers, 17.8% physically inactive, and 4.9% had a diabetes diagnosis, 56.1% hypertension, and 5.2% cardiovascular disease. Around 0.2% (N=760) of the sample were hospitalized with a COVID-19 infection during the follow-up period, and their risk profile was characterized as being male, older age, smokers, physically inactive, less highly educated, non-white ethnicity, and higher prevalence of cardiometabolic comorbidity (Table 1).

**Table 1.** Baseline characteristics of sample in relation to COVID-19.

|  | COVID-19 hospitalisation |  |
| --- | --- | --- |
|  | No | Yes |
| Age (yrs) | 56.4 ± 8.0 | 57.1 ± 9.0 |
| Sex (% men) | 44.8 | 55.3 |
| Smokers | 9.8 | 11.9 |
| Physical inactivity | 17.8 | 25.0 |
| Moderate alcohol intake | 36.2 | 28.6 |
| Degree educated | 32.8 | 26.7 |
| White ethnicity | 94.5 | 86.7 |
| Diabetes | 4.8 | 9.5 |
| Hypertension | 56.1 | 63.9 |
| Cardiovascular disease | 5.2 | 9.4 |
| Body mass index (kg/m<sup>2</sup>) | 27.3 ± 4.7 | 29.0 ± 5.4 |
| Waist-Hip ratio | 0.87 ± 0.1 | 0.91 ± 0.1 |
| Total cholesterol (mmol/l) | 5.7±1.1 | 5.4±1.2 |
| HDL cholesterol (mmol/l) | 1.5±0.4 | 1.3±0.3 |
| Glycated haemoglobin (mmol/mol) | 35.9±6.5 | 38.0±8.8 |
| C-reactive protein (log units) | 0.98±0.64 | 1.12±0.68 |

Results are expressed as percentage or mean ± SD.

There was a dose-dependent association between the risk of COVID-19 with worsening lifestyle scores, such that participants in the most unfavourable category had 4-fold higher risk (RR=4.41; 95% CI, 2.52, 7.71) (Table 2). These associations were little attenuated after adjustment for covariates. Risk ratios adjusted for age, sex and mutually for each lifestyle factor were raised for physical inactivity (1.32; 1. 10, 1.58), smoking (1.42; 1.12, 1.79), obesity (2.05; 1.68, 2.49) but not for heavy alcohol consumption (1.12; 0.93, 1.35) in relation to COVID-19 compared to optimal reference categories.

**Table 2.** Combined and individual lifestyle behavioral risk factors in relation to COVID-19 hospitalisation (N=387,109)

| Total lifestyle score | CASES/N | Relative Risk (95% CI) |  |
| --- | --- | --- | --- |
|  |  | Model 1 | Model 2 |
| 0 (optimal) | 13 / 19,776 | 1.0 (ref) | 1.0 (ref) |
| 1 | 55 / 52,053 | 1.58 (0.86, 2.59) | 1.48 (0.81, 2.71) |
| 2 | 142/ 77,861 | 2.73 (1.55, 4.81) | 2.43 (1.38, 4.29) |
| 3 | 163/ 87,998 | 2.76 (1.57, 4.85) | 2.41 (1.37, 4.25) |
| 4 | 160/ 75,123 | 3.12 (1.77, 5.49) | 2.70 (1.53, 4.75) |
| ≥5 (worst) | 227/74,298 | 4.41 (2.52, 7.71) | 3.73 (2.12, 6.54) |
| p-trend |  | <0.001 | <0.001 |
| **Individual behaviours** |  |  |  |
| *Smoking* |  |  |  |
| Never | 354/214,828 | 1.0 (ref) | 1.0 (ref) |
| Past | 313/134,855 | 1.34 (1.15, 1.56) | 1.36 (1.15, 1.59) |
| Current | 93/37,426 | 1.45 (1.16, 1.83) | 1.36 (1.08, 1.71) |
| *Physical activity* |  |  |  |
| Sufficient | 382/209,489 | 1.0 (ref) | 1.0 (ref) |
| Insufficient | 192/ 108,707 | 0.98 (0.83, 1.17) | 0.99 (0.84, 1.18) |
| None | 186/68,913 | 1.51 (1.27, 1.81) | 1.38 (1.15, 1.64) |
| *Alcohol consumption* |  |  |  |
| Below guideline | 216/140,908 | 1.0 (ref) | 1.0 (ref) |
| Rarely/never | 304/116,389 | 1.88 (1.55, 2.24) | 1.57 (1.31, 1.88) |
| Above guideline | 240/129,812 | 1.23 (1.00, 1.45) | 1.24 (1.03, 1.50) |
| *Body mass index* |  |  |  |
| Healthy weight | 166/131,162 | 1.0 (ref) | 1.0 (ref) |
| Overweight | 317/165,052 | 1.41 (1.16, 1.70) | 1.32 (1.09, 1.60) |
| Obesity | 277/90,895 | 2.28 (1.88, 2.77) | 1.97 (1.61, 2.42) |

Model 1 adjusted for age and sex
Model 2 adjusted for age, sex, education, ethnicity, diabetes, hypertension, cardiovascular disease (heart attack, angina, or stroke)

Using the Health Survey for England prevalence estimates (17% for current smoking, 25% for ex-smoking, 27% for physical inactivity, 35% for overweight and 28% for obesity), the PAF for the three unhealthy lifestyle factors in combination was 51.4% (13.3% for smoking, 8.6% for physical inactivity, and 29.5% for overweight and obesity).

## Discussion

The present study demonstrates associations between adverse lifestyle and higher risk of COVID-19 in a large community-dwelling cohort. The associations were not explained by taking into account covariates such as education, ethnicity and self-reported cardiometabolic diseases. Based on UK risk factor prevalence estimates, unhealthy behaviors in combination accounted for up to 51% of the population attributable fraction of severe COVID-19.

There are several caveats to our work. Some cases of COVID-19 could have been captured in patients originally hospitalized for reasons other than the infection. We did not capture COVID-19 infections treated outside hospital settings; rather, our outcome was people with the infection of sufficient severity to warrant in-patient care. PAF reflects the prevalence of the risk factor in the population and the strength of its association with the outcome being considered; the core assumption is that the risk factor has a causal link to the outcome. As our results are based on observational data rather than an intervention, the present PAF-findings may overestimate of the proportion of COVID-19 hospitalisation that could be been prevented by lifestyle change.

In conclusion, these data suggest that adopting simple lifestyle changes could lower the risk of severe COVID-19 infection.

## Data Availability

This research has been conducted using the UK Biobank Resource under Application 10279.

http://www.ukbiobank.ac.uk/

## Acknowledgements

Funding: GDB is supported by the UK Medical Research Council (MR/P023444/1) and the US National Institute on Aging (1R56AG052519-01; 1R01AG052519-01A1); MK by the UK Medical Research Council (MR/R024227), US National Institute on Aging (NIH), US (R01AG056477), NordForsk (75021), and Academy of Finland (311492); There was no direct financial or material support for the work reported in the manuscript.

Acknowledgement: We thank the UK Biobank study member for their generosity in participating.

Access to data: This research has been conducted using the UK Biobank Resource under Application 10279. (http://www.ukbiobank.ac.uk/)

## Contributions

MH and GDB generated the idea for the present paper, and with MK formulated an analytical plan; CRG prepared the data set; MH carried out all the data analyses and wrote the manuscript; All authors commented on an earlier version of the manuscript. MH will act as guarantors for this work. The corresponding author attests that all listed authors meet authorship criteria and that no others meeting the criteria have been omitted.

